# College Openings in the United States Increased Mobility and COVID-19 Incidence

**DOI:** 10.1101/2020.09.22.20196048

**Authors:** Martin S. Andersen, Ana I. Bento, Anirban Basu, Christopher R. Marsicano, Kosali I. Simon

## Abstract

School and college reopening-closure policies are considered one of the most promising non-pharmaceutical interventions for mitigating infectious diseases. Nonetheless, the effectiveness of these policies is still debated, largely due to the lack of empirical evidence on behavior during implementation. We examined U.S. college reopenings’ association with changes in human mobility within campuses and in COVID-19 incidence in the counties of the campuses over a twenty-week period around college reopenings in the Fall of 2020. We used an integrative framework, with a difference-in-differences design comparing areas with a college campus, before and after reopening, to areas without a campus and a Bayesian approach to estimate the daily reproductive number (*R*_*t*_). We found that college reopenings were associated with increased campus mobility, and increased COVID-19 incidence by 3.4 cases per 100,000 (95% confidence interval [CI]: 1.5 – 5.4), or a 25% increase relative to the pre-period mean. This reflected our estimate of increased transmission locally after reopening. A greater increase in county COVID-19 incidence resulted from campuses that drew students from counties with high COVID-19 incidence in the weeks before reopening (*χ*^2^(2) = 10.19, *p* = 0.006). Even by Fall of 2021, large shares of populations remained unvaccinated, increasing the relevance of understanding non-pharmaceutical decisions over an extended period of a pandemic. Our study sheds light on movement and social mixing patterns during the closure-reopening of colleges during a public health threat, and offers strategic instruments for benefit-cost analyses of school reopening/closure policies.

---

One of the key lessons learned from the COVID-19 pandemic has been the pivotal role of human behavior, specifically mobility and mixing in spreading infection, and the role of young adults. In the United States and globally, these phenomena are acutely important in congregate and communal living settings that are common not only in colleges and prisons but also in nursing homes^1–4^. However, the role of communal living, and its interaction with mobility and mixing, is difficult to identify empirically since people enter communal living settings non-randomly. The resumption of teaching on a college campus provides a sudden change in a community’s exposure to communal living and differences across college campuses lead to variation in the extent to which campus reopenings induce mixing with higher and lower incidence areas.

The susceptibility of children and college-age individuals to COVID-19 and their role in transmission has been heavily debated and remains hard to quantify^5–9^. Following the first wave of school closures in the United States in the spring of 2020, COVID-19 incidence fell across the country, leading many public health officials to view closing schools as a viable strategy to mitigate the spread of the pandemic^10,11^. However, closing schools, while potentially reducing transmission, may adversely affect children and college students. As a result, it is important to understand what role college reopenings play, if any, in the COVID-19 pandemic to design efficient mitigation strategies now and in the future.

During late Summer 2020, colleges and universities across the United States reopened and welcomed hundreds of thousands of students back to campus in the United States^12^. Over half of these institutions reopened for in-person teaching, although many institutions switched to online instruction after rapid increases in reported COVID-19 cases on campuses and in the community^13,14^. A few studies have sought to formally test the hypothesis that reopening college campuses increased COVID-19 incidence^4,15–18^. However, the institutions in these studies represent a small proportion of the 11 million undergraduates enrolled in public and non-profit four-year institutions across the country^12^. A phylogenetic study from western Wisconsin^3^ identified two clusters of SARS-CoV-2 strains on college campuses that may have subsequently infected nursing home residents, demonstrating transmission between college campuses and the surrounding community. Nonetheless, the effectiveness of college reopening policies as non-pharmaceutical interventions for mitigating the burden of COVID-19 is still disputed. Simulation-based studies have been unable to provide public health officials with conclusive recommendations, despite detailed COVID-19 transmission datasets^19,20^). The lack of a clear direction is mostly due to insufficient data about the college-specific details and how to harness movement as proxies for behavior and mixing patterns of the population while such strategies are in place. As we approach Fall 2021, with expected mass movement events in the US, millions of college and university students will return to residential instruction. This leaves little time to achieve high levels of full vaccination necessary to prevent outbreaks. Furthermore, due to new variants now circulating, there is an increased risk of breakthrough infections^21^. Thus, it is more important than ever to understand school reopenings’ effects and mass mobility events on COVID-19 incidence.

We harnessed comprehensive, national data covering the start date and instructional method of most four-year U.S. colleges and universities together with a highly resolved dataset (both spatially and by age) from the CDC,^22^ which provided detailed demographic information on COVID-19 cases around the country. This gave us the ability to directly measure the variation in human movement patterns caused by the policy and, in addition, allowed us to identify college-age cases and assign cases based on symptom onset. We hypothesized that reopening colleges would increase COVID-19 transmission within the college community with potential spillover effects onto the neighboring populations. We also hypothesized that increases in incidence would be greater on campuses that attract students from areas with a higher incidence of COVID-19 and that these effects would be concentrated among campuses providing face-to-face instruction. While there is some compelling research around testing^23,24^ and limiting student mobility^9^ as COVID-19 mitigation strategies, it is outside of the scope of our study to understand the impact of specific actions colleges may have taken in response to rising rates.

We use an integrative framework, with a difference-in-differences design comparing areas with a college campus, before and after reopening, to areas without a campus and a Bayesian approach to estimate the daily reproductive number (*R*_*t*_). We unequivocally demonstrate that there was a marked increase in COVID-19 incidence among college-age students following the reopening of campuses. Finally, while COVID-19 case counts have been a focus of several studies, our data also allowed us to examine other public health outcomes, such as hospitalizations or deaths. Our results provide evidence of the COVID-19 impact of colleges-reopening policies locally and in neighboring areas and undoubtedly informs future events in these settings.

## Results

Our study period ran from July 5^th^ 2020 to November 1^st^ 2020, which spanned the four weeks before the first campus reopening and four weeks after the last campus reopened. Of the 3,142 counties of the United States, 784 contained a college campus from our universe of 1,360 colleges. However, over 238.0 million people live in counties with a college campus. Figure 1 of the Supplementary Information maps the campuses in our sample by teaching modality.

**Figure 1.**
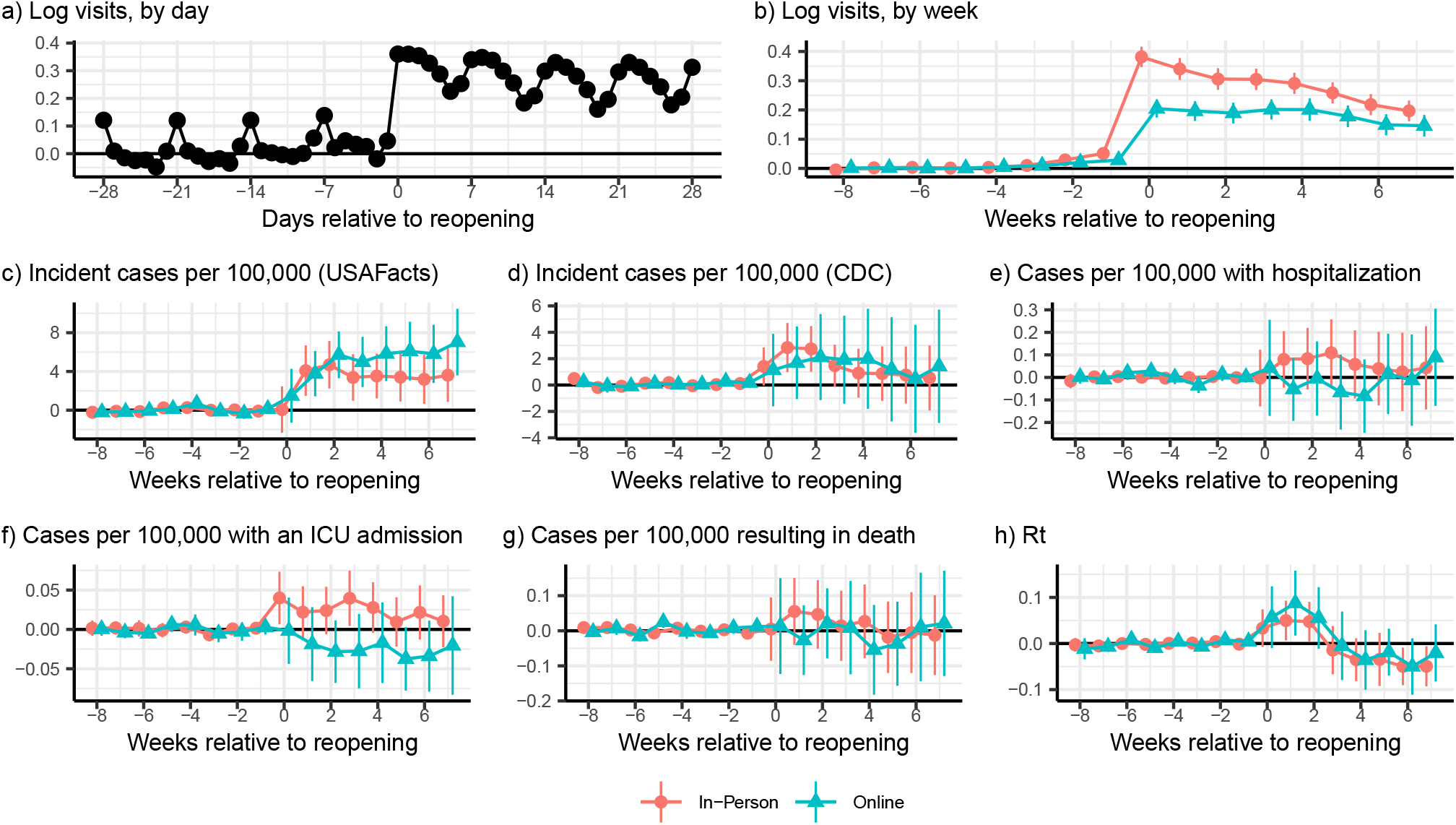
Event study estimates of reopening college campuses, relative to counties without a college campus. (a) Reopening college campuses significantly increased the number of devices on college campuses (*χ*^2^(28) = 1360.62, *p <* 0.001). These increases persisted on college campuses (b) for at least eight weeks after reopening (In-person: *χ*^2^(8) = 688.3, *p <* 0.001; Online: *χ*^2^(8) = 224.7, *p <* 0.001) and were larger for campuses that reopened for primarily in-person teaching (*χ*^2^(8) = 91.09, *p <* 0.001). Reopening college campuses also increased the incidence of COVID-19 in the county, regardless of teaching modality or data source (c and d; *χ*^2^ tests reported in the Supplementary Information). After reopening there were significantly more cases resulting in hospitalization associated with in-person, as opposed to online, reopening (*χ*^2^(8) = 16.85, *p* = 0.030). However, there were no significant differences in ICU utilization (*χ*^2^(8) = 10.74, *p* = 0.217) in the first eight weeks after reopening. During the first four weeks, in-person teaching was associated with a greater incidence of mortality, relative to online teaching (*χ*^2^(4) = 9.56, *p* = 0.049), although there were also differences in the pre-period (*χ*^2^(5) = 11.25, *p* = 0.047). Local transmission (h), measured by *R*_*t*_, was significantly different from zero after reopening a college, regardless of teaching modality (In-person *χ*^2^(8) = 36.68, *p <* 0.001; Online *χ*^2^(8) = 36.32, *p <* 0.001). COVID-19 related data are from the CDC unless otherwise specified.

Our identification strategy made comparisons between counties with and without a college campus around the time that a campus reopened in a “difference-in-differences” design^25^. Since several counties contained more than one college campus, we assigned county status based on the status of the first, and, as a tie-breaker, largest, campus to reopen in each county.

### Event studies

The reopening of a college affected mixing patterns not only of the students but also the members of the surrounding communities where these students live. The number of devices on campus increased significantly in the week before campuses reopened and remained high for at least the first 28 days following reopening (*χ*^2^(14) = 1360.62, *p <* 0.001, Figure 1a). Aggregating by week, which smooths out day of the week fluctuations in movement, and separating the sample by teaching modality demonstrated that there were significant increases in movement to census block groups containing college campuses after those campuses reopened regardless of the teaching modality (In-person: *χ*^2^(8) = 583.99, *p <* 0.001; Online: *χ*^2^(8) = 181.25, *p <* 0.001; Figure 1b), although the increase was larger for in-person reopenings (*χ*^2^(8) = 91.09, *p <* 0.001). The increase in mobility was accompanied by a rise in COVID-19 incidence, regardless of teaching modality, using national data from USAFacts (In-person *χ*^2^(8) = 39.62, *p <* 0.001; Online *χ*^2^(8) = 29.47, *p <* 0.001Figure 1c), while in-person reopenings were accompanied by increasing disease incidence using CDC data (In-person *χ*^2^(8) = 19.22, *p* = 0.014; Online *χ*^2^(8) = 4.88, *p* = 0.770 Figure 1d). The increase in COVID-19 cases was not accompanied by increases in cases requiring hospitalization (In-person *χ*^2^(8) = 11.81, *p* = 0.160; Online *χ*^2^(8) = 14.52, *p* = 0.069, Figure 1e) and that resulted in death (In-person *χ*^2^(8) = 7.67, *p* = 0.466; Online *χ*^2^(8) = 12.27, *p* = 0.140, Figure 1g). *R*_*t*_ increased during the first four weeks regardless of the teaching modality chosen by the college (In-person *χ*^2^(4) = 17.33, *p* = 0.002; Online *χ*^2^(4) = 15.33, *p* = 0.004, Figure 1h).

The weekly event study coefficients are presented in table 8 of the Supplementary Information.

### Difference-in-differences estimates

The reopenings lead to a cascade of indirect effects at the population level. To show this, we estimated a series of difference-in-difference models to estimate the effect of reopening a college campus on mobility and COVID-19 outcomes. We present the detailed results in the Supplementary Information (Table 2), but describe the results below.

Reopening a college campus was associated with a 27.8 (95% CI: 25.3-30.4, *p <* 0.001) log point increase in the number of devices on campus, or approximately a 32.0% (95% CI: 28.8-35.5%) increase in movement on campus, from the week prior to the start of classes (Table 2, column (1)). While we do find evidence of differential pre-trends (*χ*^2^(5) = 155.47, *p <* 0.001), we can bound the effect of these trends, assuming that they are linear, in the post-period which yields statistically significant effects of college reopenings on mobility (see the Supplementary Information for details). The increase in movement was larger in schools that reopened for primarily in-person, as opposed to primarily online, instruction (33.3 [29.7-36.9] vs. 19.8 [16.6 - 23.0] log points; *χ*^2^(1) = 30.07, *p <* 0.001). The increase in mobility was also larger for colleges that had greater exposure to students from areas with high levels of COVID-19 incidence (*χ*^2^(2) = 11.37, *p* = 0.003).

Using our difference-in-difference framework, we found that reopening a college was associated with a statistically significant increase of 3.4 cases per 100,000 people (95% CI: 1.5-5.4, *p <* 0.001; Table 2, column (2)) using case data from USAFacts (or approximately 25% relative to the pre-period mean). The estimate was similar, although slightly smaller, using data from the CDC sample of counties (2.0 cases per 100,000 95% CI: 0.6-3.5, *p* = 0.006; Table 2, column (3); 16% of the pre-period mean). In both cases, the results were robust to allowing for linear post-trends. The increase in COVID-19 incidence, using USAFacts data, was significantly different across terciles of exposure to COVID-19 due to student mobility (*χ*^2^(2) = 10.19, *p* = 0.006) with increased incidence in the first (*p* = 0.041) and second (*p <* 0.001) terciles.

Using data from the CDC, we found that reopening a college campus for in-person teaching increased the incidence of cases requiring an ICU admission by 0.031 (95% CI: 0.002-0.060, *p* = 0.035) cases per 100,000 (Table 2, column (4)), or almost 40%, with no effect among campuses that reopened primarily online (*p* = 0.371).

The central epidemiological parameter governing a disease system’s dynamics is the effective reproduction number (*R*_*t*_). We estimated a significant increase in daily (*R*_*t*_) around the time of reopening, consistent with an uptick in transmission. On average there was an increase in *R*_*t*_ of 0.034 (95% CI: 0.001 - 0.068, *p* = 0.043). We note that *R*_*t*_ did not significantly differ by the teaching method chosen for the campus (*χ*^2^(1) = 0.34, *p* = 0.562; Table 2) but did decline over time (*χ*^2^(2) = 39.27, *p <* 0.001).

### Age-specific incidence

To observe the age-stratified dynamics, we explored age-specific incidence. Our analyses supported the conclusion that the shifts in age dynamics overtime likely resulted from college reopenings in Figure 2. The top panel (Figure 2a) demonstrates a clear shift, where we observe an increase in COVID-19 incidence in people ages 10 - 29, but not for any other age group (*χ*^2^(5) = 47.26, *p <* 0.001). However, the increase in these college-aged students was dramatic, with incidence among 10-19 year-olds rising by 7.1 (95% CI: 4.5-9.7, *p <* 0.001) cases per 100,000, or almost 70% of the pre-period mean, while for 20-29 year-olds the increase was 6.2 (95% CI: 3.1-9.2, *p <* 0.001) cases per 100,000 (32% of the pre-period mean). The second panel indicates that our estimates of the effect of reopening on hospitalizations by age group are too noisy to draw any inferences (*χ*^2^(5) = 5.98, *p* = 0.308). The third and fourth panels demonstrate that there was a statistically significant age-specific increase in cases requiring an ICU admission among people 70 and older (0.132, 95% CI: 0.016 - 0.247, *p* = 0.025; 44% of the per-period mean), although we fail to reject the null that the effect of reopening is equal across age groups (*χ*^2^(5) = 6.73, *p* = 0.242). There are no statistically significant differences in the age-profile of COVID-19 related mortality following a campus reopening.

**Figure 2.**
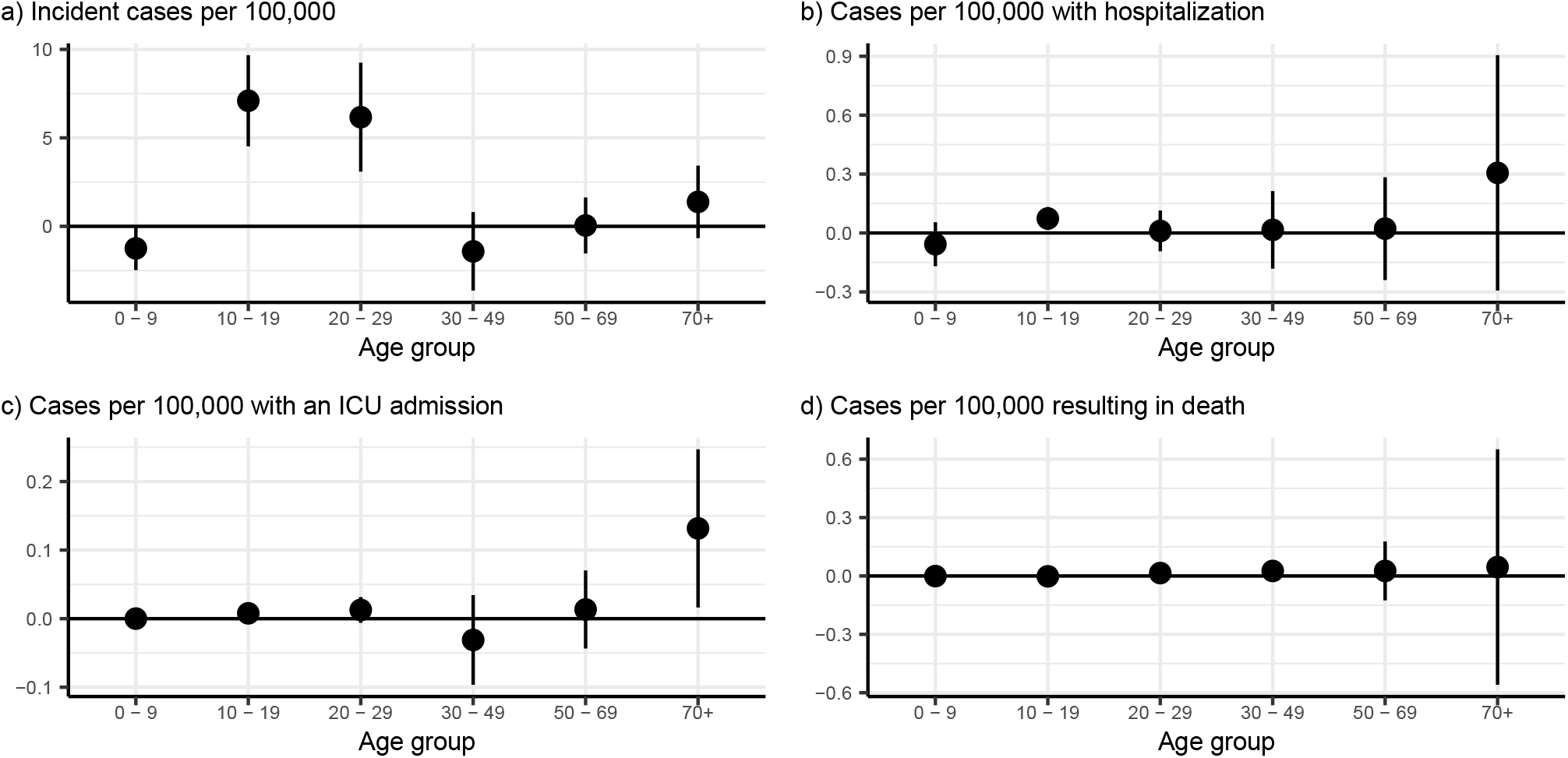
Age-specific effects of college reopenings. (a) demonstrates that the increase in the incidence of COVID-19 was isolated to people between 10 and 29 years of age, which encompasses most college-age individuals (*χ*^2^(5) = 47.26, *p <* 0.001). In the aggregate, hospitalization rates did not change differentially by age after reopening (b; *χ*^2^(5) = 5.98, *p* = 0.308), although there was a significant increase among 10-19 year-olds (0.074, 95% CI: 0.015 - 0.132; *p* = 0.014). Similarly, we find no evidence of differential changes in the incidence of cases requiring ICU admission (c; *χ*^2^(5) = 6.73, *p* = 0.242). We did not find any age-specific increases in mortality due to COVID-19, although these results were imprecisely estimated (d). Figure plots point estimates and 95% confidence intervals. Point estimates and standard errors are available in Table 3 of the Supplementary Information.

While the data appear to paint a clear picture, it is possible that several mechanisms may yield a similar age-specific profile of cases. Thus, in the SI, we test these observations. We show age-specific event studies (Figure 4) for our four age-specific outcomes. These event studies clearly demonstrate that all age groups were trending similarly prior to the reopening, except for incidence (cases: *χ*^2^(35) = 50.18, *p* = 0.046; hospitalizations: *χ*^2^(35) = 43.86, *p* = 0.145; ICU *χ*^2^(35) = 48.94, *p* = 0.059; and deaths (*χ*^2^(35) = 38.66, *p* = 0.301). COVID-19 incidence rose beginning in the week campuses reopened and remained elevated subsequently, with statistically significant increases for at least four weeks after reopening (*χ*^2^(20) = 61.86, *p <* 0.001). The event studies for hospitalization, ICU admissions, and death typically present little evidence of differences by age group either before, or after, reopening.

### Differential effects by teaching modality

We disaggregated teaching modality into more granular categories to explore potential differences across teaching methods. Our results demonstrate that campuses that reopened with a greater emphasis on in-person teaching were associated with larger increases in mobility (*χ*^2^(4) = 79.99, *p <* 0.001) and incident COVID-19 cases *χ*^2^(4) = 11.08, *p* = 0.026) (Figure 3). However, these results are tempered since at the extremes of “Fully in-person” and “Fully online” we rejected the null hypothesis of no differential pre-trends across counties (Table 6 of the Supplementary Information). Event studies (Figure 5 of the Supplementary Information) demonstrate that these two groups exhibited extreme behavior, relative to the other three teaching modality groups.

**Figure 3.**
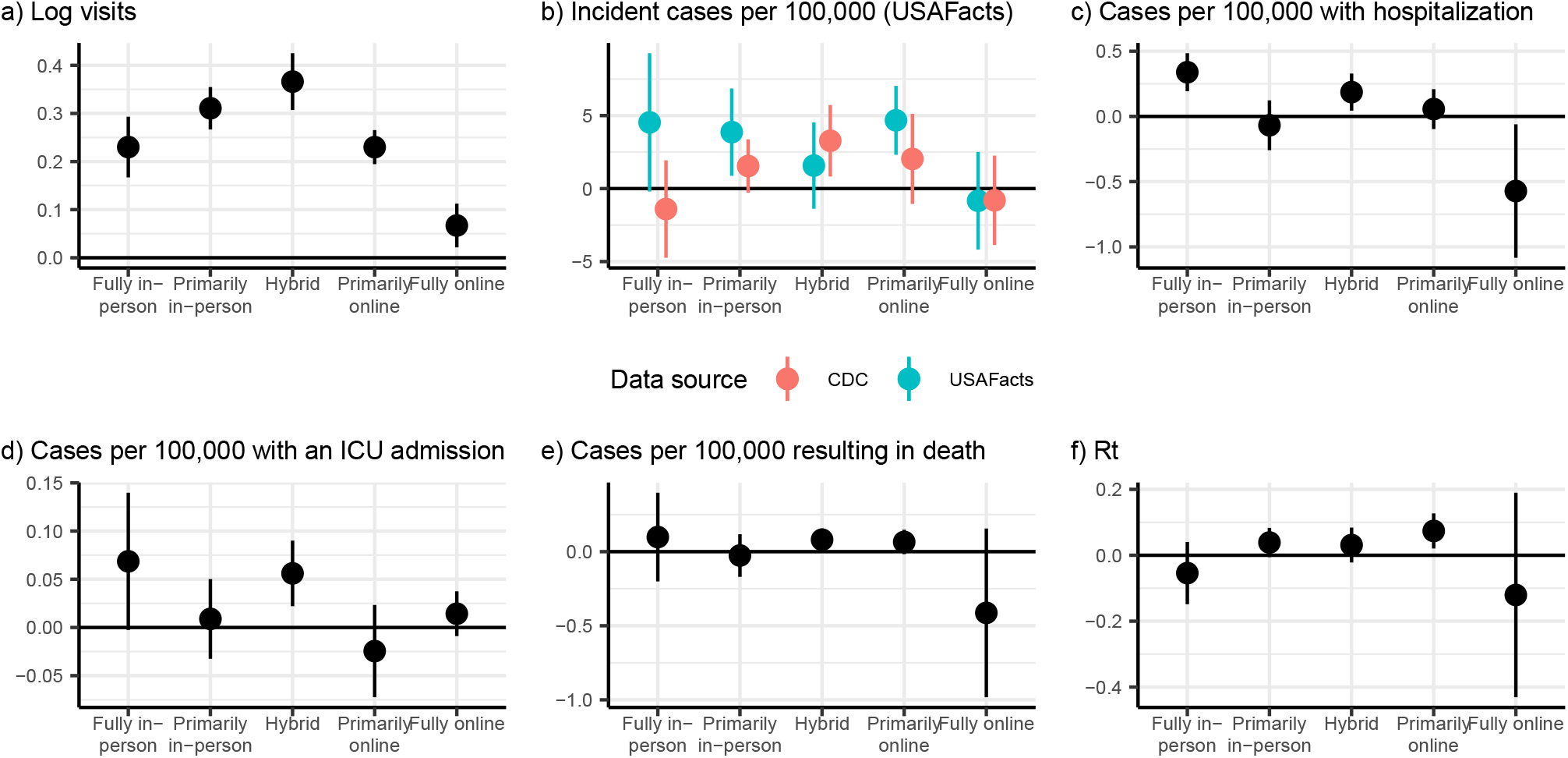
Differential effects of reopening college campuses by expanded teaching modality. (a) Campuses that reopened for “Primarily in-person” and “Hybrid” teaching had the largest increase in devices on campus following reopening, while the increase in visitors was significantly smaller for fully online reopenings (*χ*^2^(4) = 79.99, *p <* 0.001). All reopenings except “Fully Online” were associated with a significant increase in COVID-19 cases after reopening (b) using incidence data from USAFacts (*χ*^2^(4) = 11.08, *p* = 0.026). There were no statistically significant adverse effects of reopening a college campus by teaching modality for hospitalizations (c) or mortality following a reopening (e), although there were significant increases in ICU admissions (d; *χ*^2^(4) = 10.96, *p* = 0.027). Except at the extremes, for which we had relatively few colleges, there were uniform increases in *R*_*t*_, regardless of the teaching modality (f).

## Discussion

Schools and universities welcomed millions of students back to campus for Fall 2021 while vaccine uptake stagnated^26^ and many people on campuses and in surrounding communities remain vaccine hesitant. Therefore, revisiting the effects of mass movement events into campuses and how they affect local and surrounding communities is crucial. Our results provide a quantitative evaluation of mobility patterns during periods of reactive college closure and reopening strategies and highlight their impact in shaping social interactions of the college and surrounding communities. We found that college policies induced marked changes in the overall number of daily mobility interactions. Our findings demonstrate that re-opening a college was associated with an increase in the number of cellular devices on campus (a dramatic increase in population size) after classes resumed for all teaching modalities, although the increase in mobility is larger for in-person as opposed to online teaching. We unequivocally showed that re-opening a college significantly increased the incidence of COVID-19 in the county. In counties the reopened for in-person teaching teaching we also demonstrated that reopening increased the incidence of cases resulting in an ICU admission. We also demonstrated that counties containing colleges that drew students from areas with higher COVID-19 incidence experienced significantly larger increases in COVID-19 incidence following campus reopening. This is likely induced by the dramatic increase in the number of contacts of students with each other on the campuses and with the surrounding communities.

To contextualize our findings, there are 238.0 million Americans in the 786 counties that contain a college campus in our sample. Our results demonstrate that reopening college campuses resulted in an additional 8,000 (8137 [95% CI: 3515 – 12759]) cases of COVID-19 per day. This estimate is consistent with the aggregate number of cases reported on the New York Times case tracker which reported more than 397,000 cases as of December 11 2020^27^, which would correspond to seven weeks of additional cases using our main estimate and excludes spillover effects into the community.

However, because of the nature of the cases reports data, we were unable to disentangle how many of the cases we measure as our outcome are “imported” (student arrivals) and how many are local transmissions from the students. Further, asymptomatic cases were only identified if testing was done on campus regardless of symptoms. Nevertheless, our results are inconsistent with large numbers of “imported” cases since an imported case would lead to an increase in COVID-19 cases contemporaneously with any increase in mobility, while we observed a one-week lag between peak mobility and the peak change in COVID-19 incidence, when cases are assigned based on symptom onset.

We did not quantify potential spillovers to the communities surrounding campuses, as these effects would require college-level incidence data, which are not consistently collected. However, using age-specific data, we were able to demonstrate that most of the increase in COVID-19 incidence arose among college-aged students (ages 10-29).

Additional work is necessary to identify the optimal reopening-closure policies (e.g., lengths) and under which circumstances specific policies are cost-effective. However, evaluating the effectiveness of specific mitigation measures taken by colleges, especially the ways in which colleges have reacted to the initial increases in cases with strong countermeasures, was beyond the scope of this initial study and remain priorities for future studies. Similarly, we were unable to test what has occurred once colleges change decisions, such as changing instructional modes temporarily or encouraging students to return home^9^ since these changes were reactions to rapidly increasing case counts^28^.

While we only directly demonstrate that college campuses that were more heavily exposed to COVID-19 lead to larger increases in incidence, our results also indicate that sending students home from colleges due to high COVID-19 incidence is likely to lead to increased COVID-19 incidence in students’ home communities since the same exposure mechanism would run in reverse. Public health officials have also raised these concerns, some of whom have publicly opposed closing dormitories, even after a college or university transitioned to online education^29^. Further research on the effects of sending students home is needed to understand the risks and benefits of closing residence halls.

The nature of our data limits our results. Our mobility analysis relies on observing cellular GPS signals and these devices may not always report their location. In addition, it is unlikely that devices correspond in a one-to-one manner with people since college students may have more than one device (a phone and a cell-enabled tablet) that provide data under distinct identifiers. Second, we are unable to measure cases among college students vs. others in the county community, beyond using the age of the individual. Third, our mobility measure does not take account of students who may live in off-campus housing and take classes online.

Our results demonstrate the essential role that mixing and mobility play in seeding COVID-19 in the community and the role that congregate living settings play in providing a fertile ground for COVID-19 to expand. For example, these results highlight the role that nursing homes and prisons play in the COVID-19 pandemic and complement existing research on cross-nursing home linkages and COVID-19 incidence^1^. While we expect that continued testing on college campuses and current vaccination efforts will mitigate some of the effects we observed, the rate of vaccination remains low, particularly among college-age individuals^30^, and the majority of colleges did not engage in high-quality testing regimes in the 2020-2021 academic year^31^. Combined with the continued emergence of new variants and rising incidence in several states^32^, by the time of school start in the fall of 2021, we expect to observe similar dynamics, should no other strategies be implemented.

Our analysis is a good step towards building a framework to map mobility to contacts (as COVID-19 era contact matrices become available^33^), in the analysis of airborne infectious diseases. As such, our framework has a much wider scope than the study of COVID-19 related college policies in one specific region. Our findings are critical in the context of adapting public health management strategies, as they consider additional strategies to mitigate disease burden and decrease transmission. The effects of college reopenings are also informative for outbreak management in other communal settings, including nursing homes and prisons, both of which have been particularly hard hit by the COVID-19 pandemic.

Despite all data limitations, our study, provides (i) empirical evidence about changes in “behavior” (mobility surges) of the population during the implementation of the school-reopening strategies, (ii) a multi pronged approach to estimate mobility patterns and evaluate effects on the spread of infectious diseases with an unique degree of detail and (iii) tools for evidence-based decision-making beyond evaluating college reopening strategies.

## Methods

### Study Data

#### College characteristics

We collected data on opening dates and announced instructional methods from the College Crisis Initiative at Davidson College (C2i)^34^ for 1,431 public and non-profit colleges and universities (“colleges”) in the United States. The College Crisis Initiative collects data on nearly all non-profit and public four-year degree-granting institutions with full-time undergraduates that receive Title IV aid. It excludes four-year for-profit institutions, specialty institutions like seminaries or stand-alone law schools, or institutions with graduate-only programs. This list comes from the Integrated Postsecondary Education Data System (IPEDS), which lists in total 6,527 institutions ranging from research universities to non-degree-granting institutions like local cosmetology schools. IPEDS indicates that of those, 2,009 are four-year public and non-profit degree-granting institutions with first-time, full-time undergraduates. Our sample, therefore, represents nearly 70 percent of these institutions. Further, this represents 70 percent of total undergraduate enrollment among all institutions of higher education in the United States (author calculations based on IPEDS administrative 2018 data).

We assigned college campuses to Census Block Groups (CBGs) using a college campus shapefile (geographic coordinates) prepared by the Department of Homeland Security^35^. We used a spatial join to assign each Census Block Group to the college campus that occupied the largest area in the block group, as a result, our assignment of campuses to block groups was unique. We then merged these data with college opening dates. Our final sample included 1371 schools in 786 counties. We assigned reopening strategies based on the mode of instruction reported on the date instruction began for Fall 2020. Campuses were classified as in-person or online based on the instructional modality in effect the day classes resumed for the Fall semester. Institutions that instituted primarily hybrid (379) or primarily in-person (493) modes of instruction were classified as “in person.” Institutions that offered only online classes, or for which the majority of the classes offered were online were classified as “online” (499). We assigned instructional modalities to the 786 counties with a college in our sample based on the status of the first campus to reopen in each county and, if necessary, the largest campus of those that opened on the same day. We classified 552 counties as in-person and 234 as online (Table 7 of the Supplementary Information). In the average “In-person” county, 6% of campuses reopened for online teaching, while in the average “Online” county 14% of campuses reopened for in-person teaching.

#### Mobility

We extracted cellular data from SafeGraph’s Social Distancing Metrics files. SafeGraph aggregates anonymized location data from numerous applications in order to provide insights about physical places, via the Placekey Community. To enhance privacy, SafeGraph excludes CBG information if fewer than five devices visited an establishment in a month from a given CBG. These data measure the number of devices that are detected each day in each CBG, from June 24^th^ through November 9th. SafeGraph data have been used in several recent publications^36–41^.

#### COVID-19 cases and sequelae

We used aggregate cumualtive case data at the county level from USAFacts and deidentified, case-level data from the Centers for Disease Control to estimate the incidence of COVID-19 in a county by age-group^22^. The University of North Carolina at Greensboro Institutional Review Board reviewed and approved our use of the CDC data. The CDC does not make any claims regarding the accuracy or validity of its data therefore we restricted our use of the CDC data to those counties in which the cumulative number of cases at the end of our study period was no less than 95% of the USAFacts estimate for that same county and the correlation in the rolling thirty day incidence of cases exceeded 0.95. Figure 2 in the Supplementary Information provides a map of the 1917 counties that met our inclusion criteria for the CDC data. Using the CDC data we estimated the number of cases diagnosed in each county, age-group, date cell and the number of cases that were, by March 31, 2021, hospitalized, admitted to the ICU, or resulted in death. We converted these values into values per 100,000 people in a age-county cell using population data from the 5-year American Community Survey^42^. The CDC data identifies the ultimate outcome of cases by diagnosis date, so our data indicates the number of incident cases per 100,000 people and the number that resulted in death, hospitalization, or ICU admission.

In the Supplementary Information, we describe our estimation of the effective reproductive number (*R*(*t*)) (1.1) and how we constructed our index for college exposure to other counties (1.2).

#### Data availability

All data used can be requested from SafeGraph, the CDC, and the College Crisis Initiative or are publicly available at USAFacts.org.

#### Code availability

Code to replicate our results will be made available at github.com/andersen-hecon/Colleges_COVID_19.

### Statistical Analysis

Our main analyses use a panel of counties and Census Block Groups (CBGs). In our county level analyses we identified the earliest and, if necessary, largest college or university in each county and assigned the county that college or university’s reopening modality. We estimated generalized difference-in-differences (DiD) models^43^ for mobility to campus, COVID-19 incidence, and COVID-19 cases resulting in a hospitalization, ICU admission, or death by March 31 2021. To incorporate variation across age groups, we also used age-specific data on COVID-19 incidence and cases resulting in hospitalization, ICU admission, and death. Analyses of mobility used Census Block Group (CBG) data, while for all other outcomes we used county-level data.

For each reopening date (a group) and calendar date, the generalized DiD model computes a separate estimate of the average effect of treatment on the treated (ATT) using other counties or block groups that did not have a college as controls. In these calculations, the generalized DiD embeds a propensity score estimation and an outcome regression. We model both using the natural log of the population in the county (census block group). We aggregate these ATT estimates into daily and weekly estimates of the average effect of treatment on the treated. Using the weekly aggregate we also tested for pre-trends using *χ*^2^ tests for the joint-significance of the weekly ATTs from 7 to 3 weeks before reopening. We computed an overall DiD estimate for the effect of reopening as the average of the ATTs for the period from day 0 to day 27 following reopening and computed standard errors using a clustered multiplier bootstrap^43^.

We also computed estimates for each reopening type comparing each reopening type to counties and CBGs that did not have a college. We then computed cross-type tests for the equality of the DiD coefficients using *χ*^2^ tests after bootstrapping the covariance matrix between outcomes.

We tested for an effect of exposure to students from high-incidence counties by breaking the exposure index into terciles and computing tercile specific DiD estimates. We then tested for equality across terciles using *χ*^2^ tests.

We also assessed the robustness of our results to allowing for violations of the parallel trends assumption^44^ using the HonestDiD package in R.

Means and standard deviations of our dependent variables are presented in table 1 of the Supplementary Information.

## Data Availability

We are not permitted to redistribute the data that we use in our analysis. However, it is readily available from SafeGraph and the CDC for academic researchers.

https://github.com/andersen-hecon/Colleges_COVID_19

## Acknowledgements

We would like to thank the University of North Carolina at Chapel Hill and the Research Computing group for providing computational resources and support that have contributed to these research results. Brant Callaway provided helpful advice on implementing the DiD estimator.

## Author contributions

M.A., A.I.B., A.B., C.M., and K.S. designed the research and wrote the paper. M.A. and A.I.B. performed the research and analyzed the data. C.M. contributed novel data.

## 1 Additional Methods

### 1.1 Effective reproductive number (*R*(*t*))

We used the daily cases, incubation period, and serial interval previously estimated^1^. This allowed us to estimate the effective reproduction number for each county. The effective reproduction number *R*(*t*) represents the mean number of secondary cases generated by a primary infector at time *t*^2–5^. This measure is useful to track the effectiveness of performed control measures, which aims to push it below the epidemic threshold (corresponding to *R*(*t*) = 1). *R*(*t*) incorporates factors affecting the spread of the epidemic (e.g., individual’s behavior and susceptible depletion).

To estimate *R*(*t*), we use the same methodology described previously^4–6^ to distinguish between locally acquired and imported cases. Thus, we assume that the daily number of new cases (date of symptom onset) with locally acquired infection *L*(*t*) can be approximated by a Poisson distribution

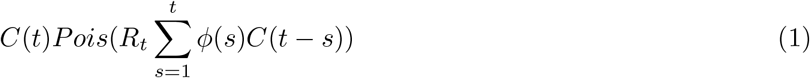

where *C*(*t*) is the number of new cases (either locally acquired or imported) at time *t* (date of symptom onset), *R*(*t*) is the effective reproduction number at time *t* and is the generation time distribution. To estimate the time between consecutive generations of cases, we adopted the serial interval (which measures the time difference between the symptom onset of the infectors and of their infected) estimated from the literature^1^, namely a gamma distribution with mean 5.0 days and standard deviation 3.4 days (shape=4.87, rate=0.65).

The likelihood *λ* of the observed time series cases from day 1 to *T* can be written as:

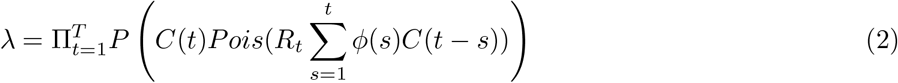

where *P* (*x, y*) is the Poisson density distribution of observing *x* events, given the parameter *y*.

We then use Metropolis-Hastings MCMC sampling to estimate the posterior distribution of *R*(*t*). The Markov chains were run for 100,000 iterations, considering a burn-in period of 10,000 steps, and assuming non-informative prior distributions of *R*(*t*) (flat distribution in the range (0-1000]). Convergence was checked by visual inspection by running multiple chains starting from different starting points.

### 1.2 Constructing exposure measures

We constructed a measure of a colleges exposure to different geographic areas using movement data from 2020. For each county and college campus we estimated the average number of devices from the source county on each campus by week and computed the change from 20 to 14 days before reopening to 0 to 6 days after reopening. Because in some cases the net flow went in the opposite direction, we truncated the change in devices at 0. We use this change in mobility as our estimate of the flow of devices from a source county to a college campus.

We then estimated, for each county, the 7-day incidence of COVID-19 per 100,000 people ending 14 days before campus reopened. Finally, we used the truncated number of devices moving from each county to a given campus to construct the weighted average of all counties that had net movement towards a college campus. The resulting exposure metric is the average 7-day incidence per 100,000 from source counties to a college campus for the period ending two weeks before campus reopened.

## 2 Robustness checks and alternative specifications

We discuss our robustness checks, which are presented in table 5 below. The first row of table 5 repeats our baseline results as a reference.

### 2.1 Restricting to counties with a college

Restricting our sample to counties with a college or university reduces the magnitude of our estimates and leaves only the increase in COVID-19 incidence resulting in hospitalization as statistically significant.

### 2.2 Allowing for linear trends

We also computed results that allow for a continuation of pre-existing linear trends, as in^7^. We report the fixed-length confidence intervals in the row labeled “Bounds for linear trends” in table 5. The first three columns demonstrate that our main results for visitors and COVID-19 incidence are robust to allowing for linear trends since the confidence intervals both contain our original point estimate and do not contain zero. The next three columns paint a more variable picture. In all cases our point estimate is contained within the robust confidence interval. However, as with our main analysis, we do not find statistically significant effects of reopening on cases requiring hospitalization, ICU admission, or death. The final column, as with the first three, demonstrates that our results are robust to allowing for linear trends in outcomes.

---

**Figure 1:**
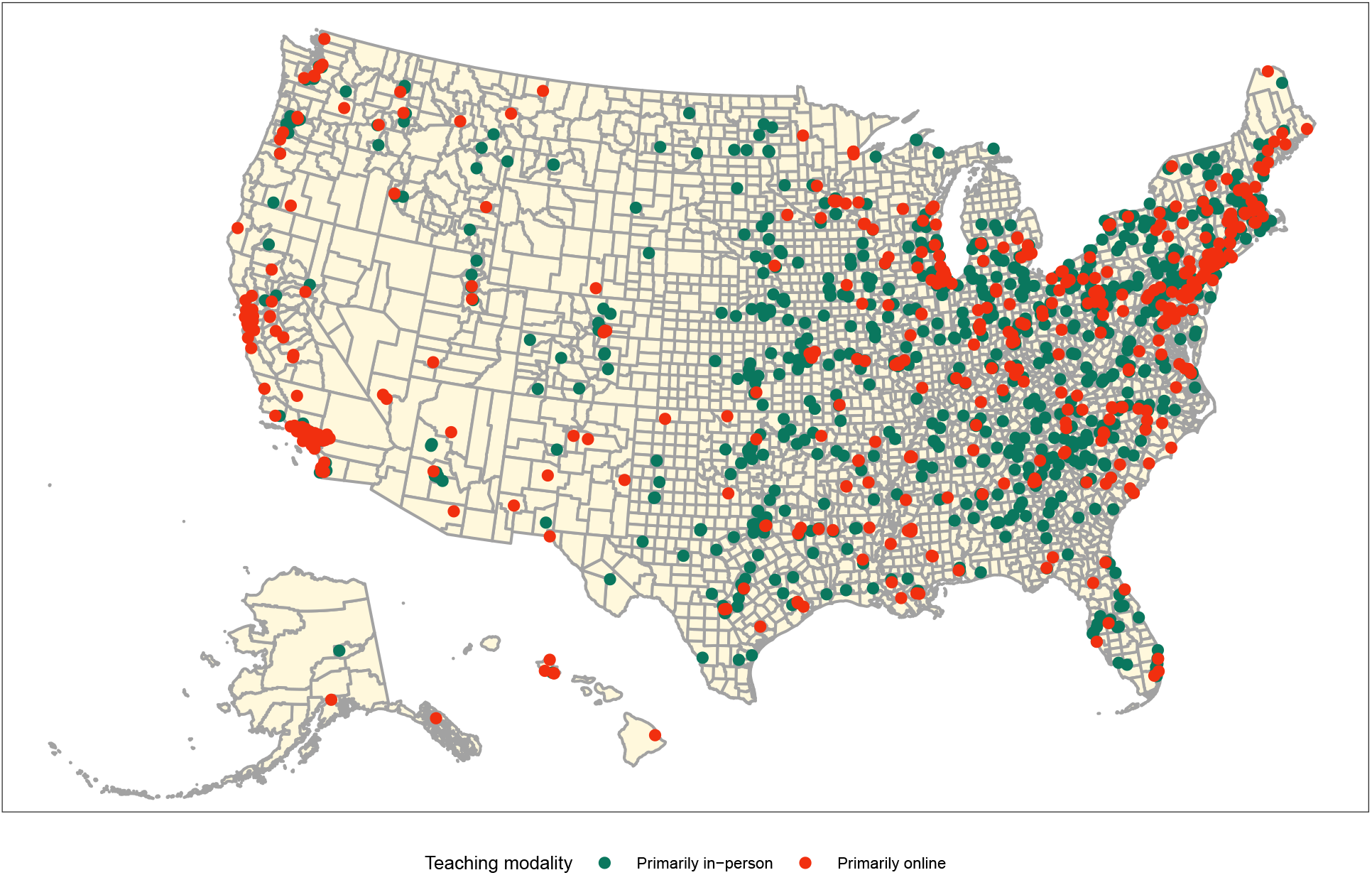
Geographic distribution of colleges and universities in sample by teaching modality. Colleges and universities are more prevalent in the eastern half of the United States, while colleges in the western half were more likely to reopen for online teaching.

**Figure 2:**
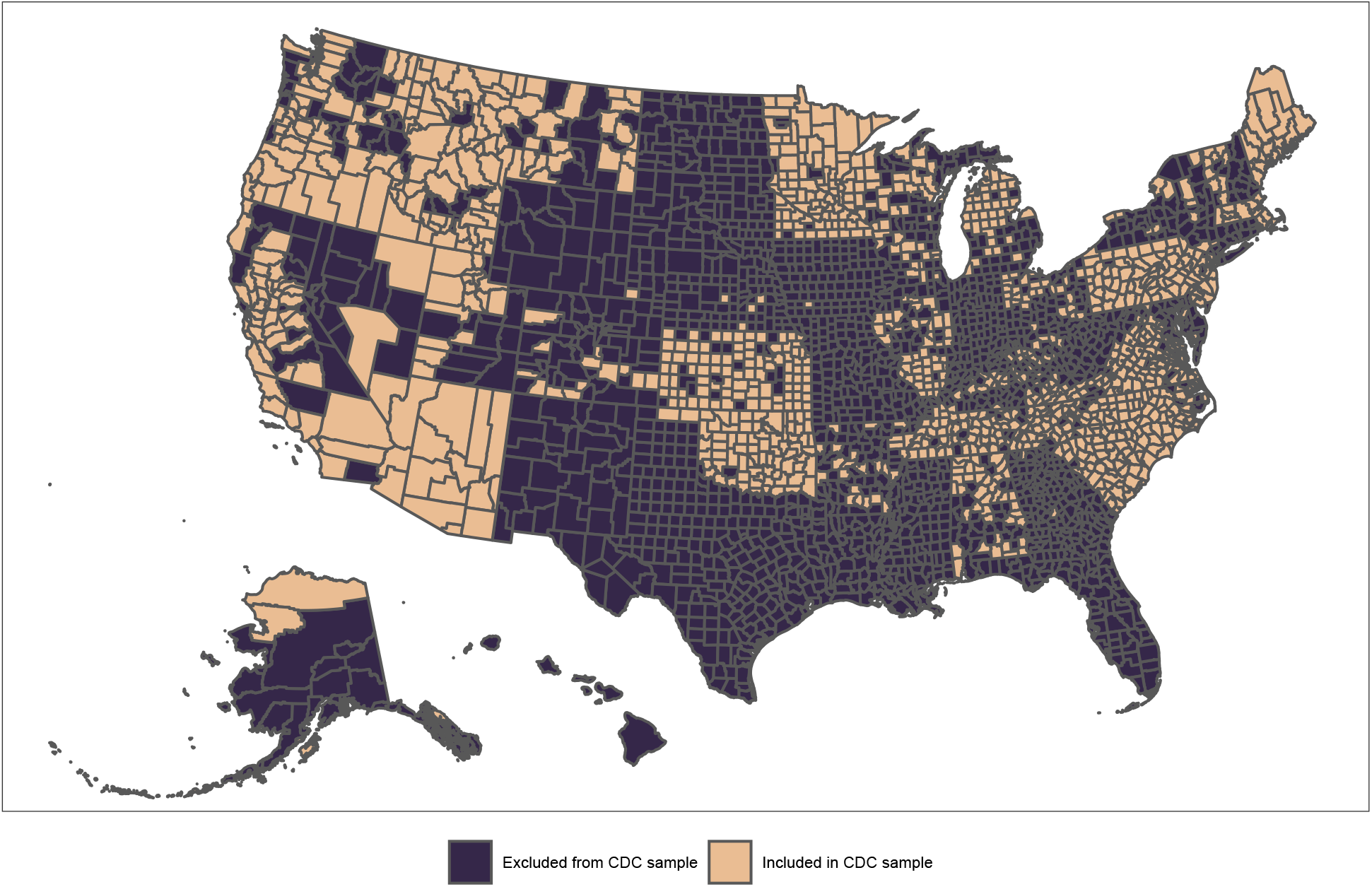
Geographic distribution of counties that are included in the CDC sample. Counties were included in the CDC sample if the ratio of cumulative cases during our study period in the CDC line files relative to USAFacts was greater than 0.95 and the correlation in the 30-day cumulative case count time series was greater than 0.95.

**Figure 3:**
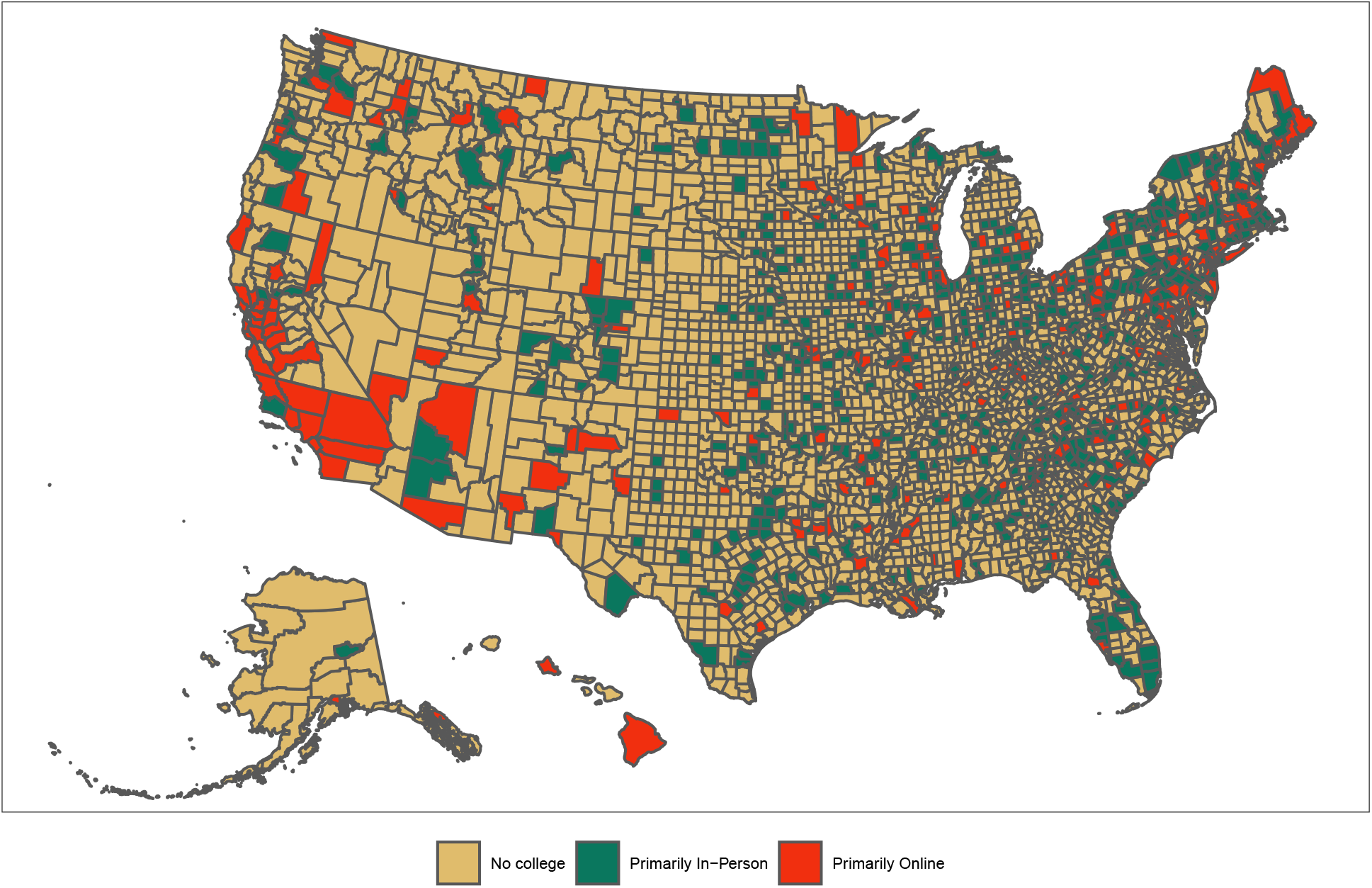
Geographic distribution of teaching assignments. Counties were assigned on the basis of the earliest and, if necessary, largest college or university in each county.

**Figure 4:**
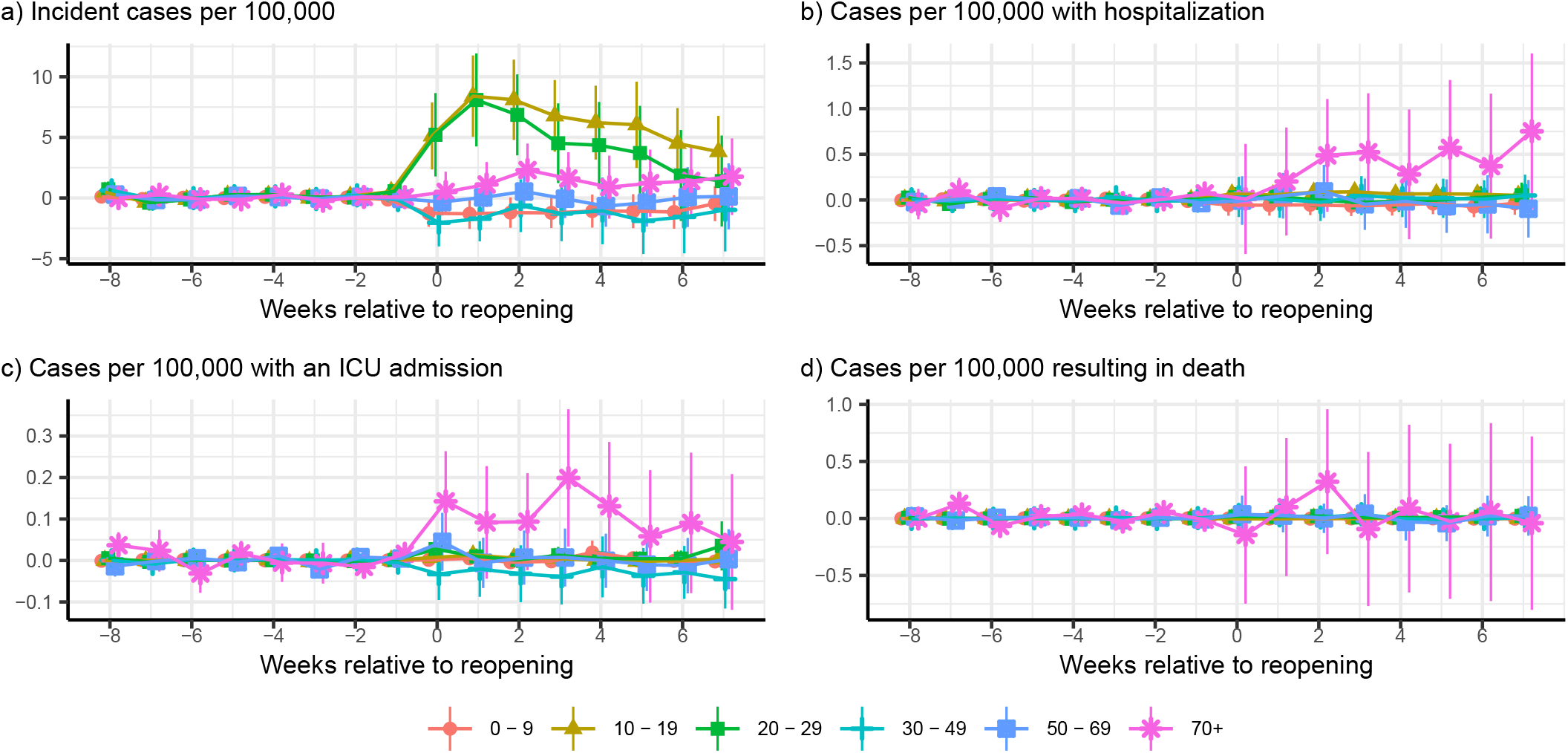
Age-specific event studies. COVID-19 incidence rose for the 10-19 and 20-29 year old age groups following the resumption of classes, although the increase faded over time for both age groups (a), while cases requiring hospitalization (b) and ICU care (c) followed less precisely estimated patterns with a decreasing time trend throughout the study period. Mortality due to COVID-19 was stable throughout the study period as well for all age groups (d).

**Figure 5:**
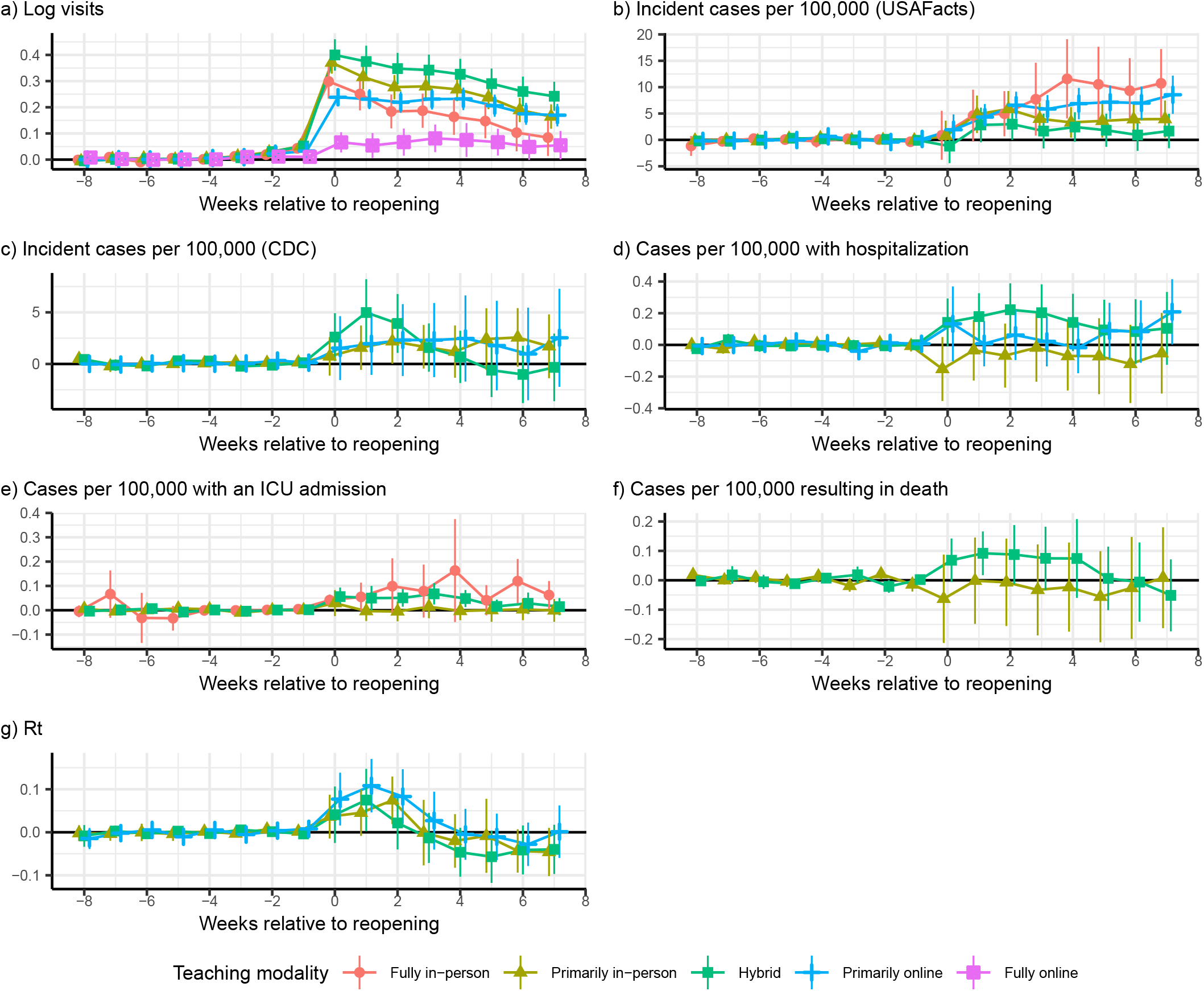
Teaching modality-specific event studies. On-campus mobility increased for all teaching modalities (a), while COVID-19 incidence rose for “Primarily in-person” (*χ*^2^(8) = 25.6, *p* = 0.001) and “Primarily online” (*χ*^2^(8) = 35.1, *p <* 0.001) teaching modalities (b and c). Hospitalization rates rose following all reopening types (d), except for primarily in-person modalities (*χ*^2^(8) = 8.9, *p* = 0.348). ICU admissions increased following reopening across for “Fully online” (*χ*^2^(8) = 40.1, *p <* 0.001) and “Hybrid” (*χ*^2^(8) = 24.8, *p* = 0.002) reopenings (e). Deaths rose following reopening for “Hybrid” (*χ*^2^(8) = 18.7, *p* = 0.016), but not “Primarily in-person” (*χ*^2^(8) = 5.2, *p* = 0.741) reopenings (f). *R*_*t*_ was significantly different following reopening for all three modalities presented (g). Except for Log visits, all panels exclude teaching modalities with statistically significant pre-trends (see Table 6 for the test statistics).

**Table 1:** Summary statistics on the sample.

|  | No college | Primarily in-person |  | Primarily online |  |
| --- | --- | --- | --- | --- | --- |
|  |  | Before | After | Before | After |
| Log visitors | 4.17 (0.558) | 4.54 (0.852) | 4.95 (0.760) | 4.27 (0.896) | 4.64 (0.870) |
| Visitors | 99.9 (62.8) | 165.0<br>(171.4) | 218.4<br>(199.7) | 134.0<br>(147.9) | 176.6<br>(178.5) |
| Daily new cases per 100,000... |  |  |  |  |  |
| ...from USAFacts | 20.7 (54.8) | 13.5 (49.6) | 21.6 (33.1) | 13.0 (19.6) | 19.1 (39.3) |
| ...from CDC | 20.9 (34.5) | 12.3 (14.9) | 21.1 (23.4) | 12.6 (14.6) | 19.4 (25.3) |
| ...resulting in hospitalization | 1.156 (3.80) | 0.742 (1.39) | 0.949 (1.68) | 0.761 (1.27) | 0.847 (1.53) |
| ...resulting in ICU admission | 0.1191<br>(1.222) | 0.0794<br>(0.415) | 0.0905<br>(0.473) | 0.0905<br>(0.308) | 0.0734<br>(0.311) |
| ...resulting in death | 0.483<br>(2.548) | 0.226<br>(0.729) | 0.375<br>(1.067) | 0.237<br>(0.861) | 0.321<br>(1.057) |
| $R_t$ | 1.39 (1.269) | 1.21 (0.637) | 1.11 (0.617) | 1.18 (0.840) | 1.10 (0.410) |
| Exposure ('0000s) | — | 1.68 (1.65) | 1.77 (1.70) | 1.33 (1.38) | 1.41 (1.44) |

Source—Authors’ analysis of C2I data on college reopening, SafeGraph mobility data, and COVID-19 case and mortality data.

**Table 2:** Difference-in-difference regressions demonstrating the effect of reopening college campuses on mobility and COVID-19 incidence and sequelae.

|  | Log visitors | Daily new cases per 100,000 from USAFacts | Daily new cases per 100,000 from CDC | Daily new hospitalized cases per 100,000 | Daily new cases in ICU per 100,000 | Daily new deaths per 100,000 | Rt |
| --- | --- | --- | --- | --- | --- | --- | --- |
| Baseline | 0.278 (21.41)<br>[0.253, 0.304]<br>{<0.001} | 3.419 (3.45)<br>[1.477, 5.361]<br>{<0.001} | 2.013 (2.74)<br>[0.572, 3.454]<br>{0.006} | 0.038 (0.70)<br>[-0.069, 0.145]<br>{0.486} | 0.015 (1.12)<br>[-0.011, 0.042]<br>{0.261} | 0.021 (0.55)<br>[-0.054, 0.097]<br>{0.583} | 0.034 (2.03)<br>[0.001, 0.068]<br>{0.043} |
| In-person | 0.333 (18.34)<br>[0.297, 0.369]<br>{<0.001} | 3.055 (2.60)<br>[0.756, 5.355]<br>{0.009} | 2.116 (2.79)<br>[0.630, 3.601]<br>{0.005} | 0.067 (1.07)<br>[-0.056, 0.191]<br>{0.285} | 0.031 (2.11)<br>[0.002, 0.060]<br>{0.035} | 0.030 (0.66)<br>[-0.060, 0.120]<br>{0.510} | 0.029 (1.62)<br>[-0.006, 0.064]<br>{0.106} |
| Online | 0.198 (12.31)<br>[0.166, 0.230]<br>{<0.001} | 3.991 (3.43)<br>[1.709, 6.273]<br>{<0.001} | 1.715 (1.19)<br>[-1.120, 4.550]<br>{0.236} | -0.020 (-0.26)<br>[-0.173, 0.133]<br>{0.797} | -0.019 (-0.89)<br>[-0.061, 0.023]<br>{0.371} | 0.004 (0.08)<br>[-0.103, 0.111]<br>{0.938} | 0.049 (1.54)<br>[-0.013, 0.111]<br>{0.123} |
| Test of In-person=Online ($\chi^2(1)$ (p-value)) | 30.07 (< 0.001) | 0.48 (0.487) | 0.06 (0.802) | 0.88 (0.348) | 4.36 (0.037) | 0.14 (0.713) | 0.34 (0.562) |
| Week(s) relative to reopening |  |  |  |  |  |  |  |
| Week before | 0.042 (22.74)<br>[0.038, 0.046]<br>{<0.001} | -0.020 (-0.19)<br>[-0.225, 0.186]<br>{0.852} | 0.129 (1.30)<br>[-0.066, 0.324]<br>{0.195} | -0.001 (-0.15)<br>[-0.019, 0.016]<br>{0.878} | 0.002 (0.81)<br>[-0.003, 0.006]<br>{0.419} | -0.001 (-0.10)<br>[-0.012, 0.011]<br>{0.918} | 0.000 (0.15)<br>[-0.005, 0.005]<br>{0.881} |
| Weeks 0-1 | 0.296 (22.71)<br>[0.270, 0.321]<br>{<0.001} | 2.336 (2.21)<br>[0.262, 4.410]<br>{0.027} | 1.898 (2.58)<br>[0.455, 3.340]<br>{0.010} | 0.022 (0.40)<br>[-0.085, 0.128]<br>{0.689} | 0.018 (1.29)<br>[-0.009, 0.045]<br>{0.195} | 0.017 (0.43)<br>[-0.061, 0.094]<br>{0.667} | 0.051 (2.82)<br>[0.015, 0.086]<br>{0.005} |
| Weeks 2-3 | 0.261 (19.13)<br>[0.234, 0.287]<br>{<0.001} | 4.502 (4.49)<br>[2.538, 6.466]<br>{<0.001} | 2.128 (2.58)<br>[0.509, 3.747]<br>{0.010} | 0.054 (0.91)<br>[-0.063, 0.171]<br>{0.362} | 0.013 (0.87)<br>[-0.016, 0.041]<br>{0.386} | 0.025 (0.62)<br>[-0.055, 0.105]<br>{0.534} | 0.018 (0.94)<br>[-0.020, 0.057]<br>{0.348} |
| Weeks 4+ | 0.212 (15.68)<br>[0.186, 0.239]<br>{<0.001} | 4.409 (3.96)<br>[2.226, 6.591]<br>{<0.001} | 1.021 (0.96)<br>[-1.068, 3.109]<br>{0.338} | 0.033 (0.48)<br>[-0.103, 0.170]<br>{0.631} | 0.005 (0.39)<br>[-0.022, 0.033]<br>{0.698} | -0.003 (-0.07)<br>[-0.093, 0.086]<br>{0.944} | -0.041 (-2.36)<br>[-0.075, -0.007]<br>{0.018} |
| Test of equality across weeks 0-1, 2-3, and 4+ ($\chi^2(2)$ (p-value)) | 140.50 (< 0.001) | 14.69 (< 0.001) | 2.86 (0.239) | 1.27 (0.529) | 1.59 (0.452) | 1.43 (0.489) | 39.27 (< 0.001) |
| Terciles of COVID-19 exposure |  |  |  |  |  |  |  |
| 1st tercile | 0.279 (15.05)<br>[0.243, 0.315]<br>{<0.001} | 1.723 (2.04)<br>[0.067, 3.378]<br>{0.041} | 0.451 (0.87)<br>[-0.567, 1.470]<br>{0.385} | 0.036 (0.53)<br>[-0.097, 0.169]<br>{0.593} | -0.008 (-0.54)<br>[-0.036, 0.020]<br>{0.586} | 0.069 (2.65)<br>[0.018, 0.120]<br>{0.008} | 0.066 (1.54)<br>[-0.018, 0.149]<br>{0.124} |
| 2nd tercile | 0.225 (13.15)<br>[0.191, 0.258]<br>{<0.001} | 5.199 (4.78)<br>[3.068, 7.330]<br>{<0.001} | 2.030 (1.94)<br>[-0.017, 4.078]<br>{0.052} | -0.009 (-0.10)<br>[-0.181, 0.164]<br>{0.921} | 0.010 (0.43)<br>[-0.036, 0.056]<br>{0.669} | -0.008 (-0.10)<br>[-0.158, 0.143]<br>{0.920} | -0.001 (-0.06)<br>[-0.041, 0.038]<br>{0.950} |
| 3rd tercile | 0.317 (13.37)<br>[0.271, 0.364]<br>{<0.001} | 2.792 (1.62)<br>[-0.582, 6.166]<br>{0.105} | 3.146 (2.07)<br>[0.173, 6.120]<br>{0.038} | 0.079 (1.01)<br>[-0.074, 0.233]<br>{0.311} | 0.037 (1.89)<br>[-0.001, 0.074]<br>{0.059} | 0.010 (0.21)<br>[-0.083, 0.103]<br>{0.831} | 0.050 (2.32)<br>[0.008, 0.092]<br>{0.020} |
| Test of equality across terciles ($\chi^2(2)$ (p-value)) | 11.37 (0.003) | 10.19 (0.006) | 4.43 (0.109) | 0.59 (0.746) | 3.96 (0.138) | 1.99 (0.369) | 4.32 (0.115) |

Source—Authors' analysis of C2I data on college reopening, SafeGraph mobility data, and COVID-19 case and mortality data.
Notes—Estimates are aggregated treatment effects from generalized difference-in-differences regressions for the first 28 days following reopening. “Primarily in-person” includes “Fully in-person” and “Hybrid”; “Primarily online” includes “Fully online”. Exposure is the 7-day COVID-19 incidence in source counties for each college two weeks prior to reopening, weighted by the change in movement to the campus from each county. Column titles indicate the dependent variable; each panel is a separate specification. Z-statistics in parentheses following point estimates, 95% confidence intervals in square brackets, and p-values in curly brackets.

**Table 3:** Age-specific difference-in-differences demonstrates that the increase in incidence of COVID-19 was isolated to people between 10 and 29 years of age, which encompasses college-age individuals

|  | Daily new cases per<br>100,000 | Daily new<br>hospitalized cases per<br>100,000 | Daily new cases in<br>ICU per 100,000 | Daily new deaths per<br>100,000 |
| --- | --- | --- | --- | --- |
| 0 - 9 | -1.247 (-1.99)<br>[-2.474, -0.021]<br>{0.046} | -0.057 (-1.00)<br>[-0.169, 0.055]<br>{0.316} | 0.000 (0.02)<br>[-0.006, 0.006]<br>{0.986} | -0.001 (-0.63)<br>[-0.003, 0.002]<br>{0.528} |
| 10 - 19 | 7.097 (5.40)<br>[4.520, 9.673]<br>{< 0.001} | 0.074 (2.47)<br>[0.015, 0.132]<br>{0.014} | 0.008 (1.26)<br>[-0.004, 0.020]<br>{0.206} | -0.002 (-2.00)<br>[-0.004, -0.000]<br>{0.046} |
| 20 - 29 | 6.169 (3.93)<br>[3.093, 9.244]<br>{< 0.001} | 0.011 (0.20)<br>[-0.093, 0.114]<br>{0.842} | 0.013 (1.33)<br>[-0.006, 0.032]<br>{0.185} | 0.015 (1.89)<br>[-0.001, 0.030]<br>{0.059} |
| 30 - 49 | -1.415 (-1.25)<br>[-3.631, 0.801]<br>{0.211} | 0.016 (0.16)<br>[-0.182, 0.213]<br>{0.874} | -0.031 (-0.93)<br>[-0.096, 0.034]<br>{0.351} | 0.026 (2.19)<br>[0.003, 0.049]<br>{0.029} |
| 50 - 69 | 0.049 (0.06)<br>[-1.533, 1.631]<br>{0.952} | 0.022 (0.16)<br>[-0.240, 0.284]<br>{0.869} | 0.013 (0.46)<br>[-0.044, 0.070]<br>{0.646} | 0.026 (0.33)<br>[-0.125, 0.177]<br>{0.738} |
| 70+ | 1.382 (1.32)<br>[-0.667, 3.431]<br>{0.186} | 0.306 (1.00)<br>[-0.293, 0.906]<br>{0.317} | 0.132 (2.24)<br>[0.016, 0.247]<br>{0.025} | 0.046 (0.15)<br>[-0.559, 0.651]<br>{0.882} |
| $\chi^2(5)$ | 47.26 (< 0.001) | 5.98 (0.308) | 6.73 (0.242) | 8.41 (0.135) |

Source—Authors’ analysis of C2I data, SafeGraph mobility data, and CDC COVID-19 case data.
Notes—Estimates are aggregated treatment effects from generalized difference-in-differences regressions for the first 28 days following reopening. Column titles indicate the class of the dependent variable and the row label indicates the age group. Z-statistics in parentheses following point estimates, 95% confidence intervals in square brackets, and p-values in curly brackets. $\chi^2$ tests for equality across age groups are reported in the bottom row.

**Table 4:**
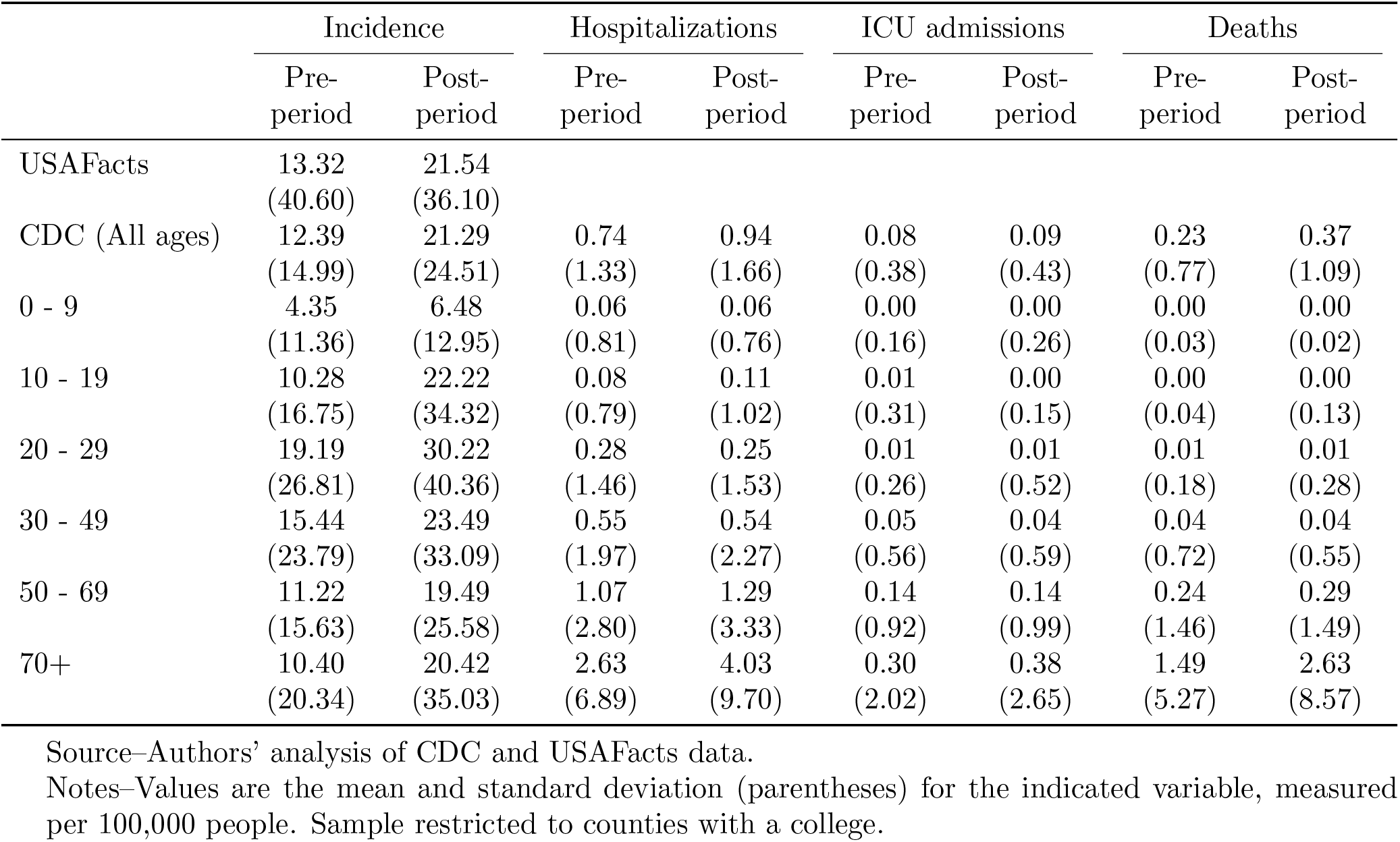
Age-specific means of the dependent variables

**Table 5:**
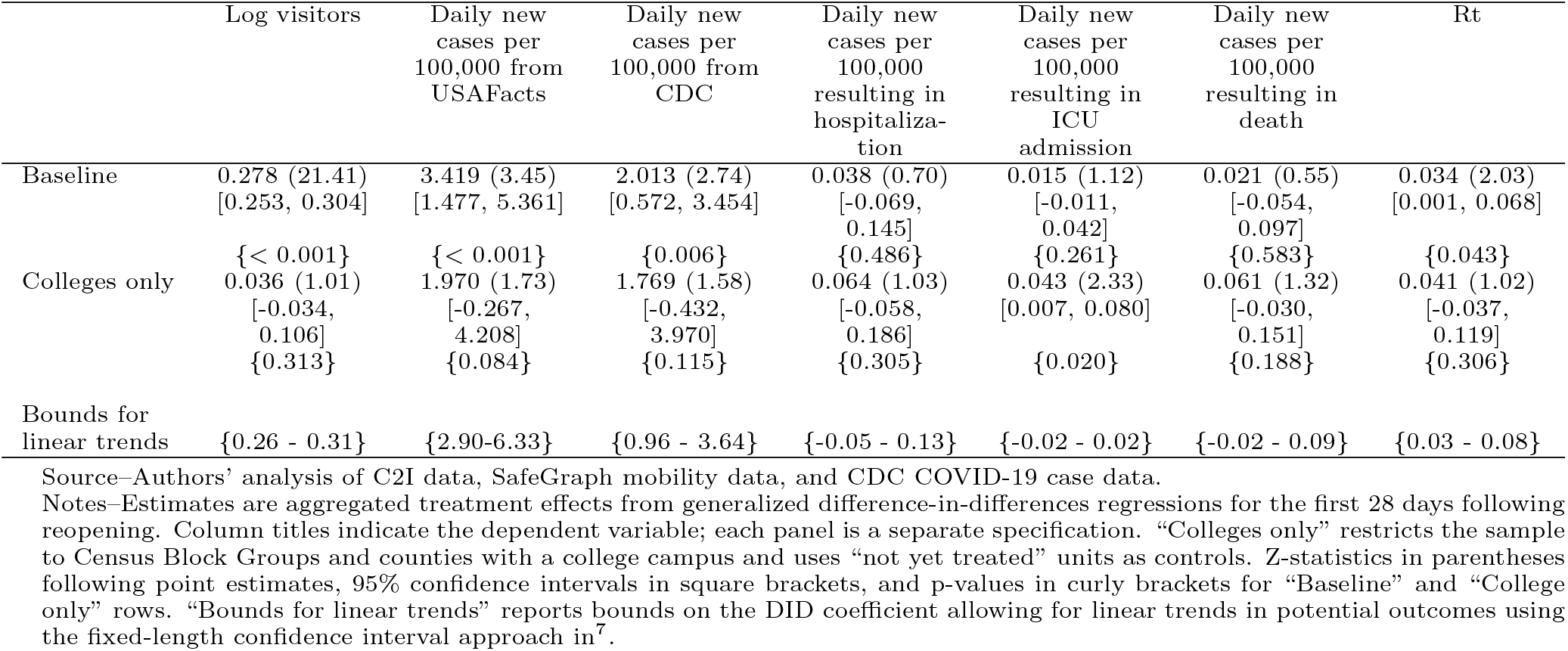
Robustness checks discussed in section 2

**Table 6:**
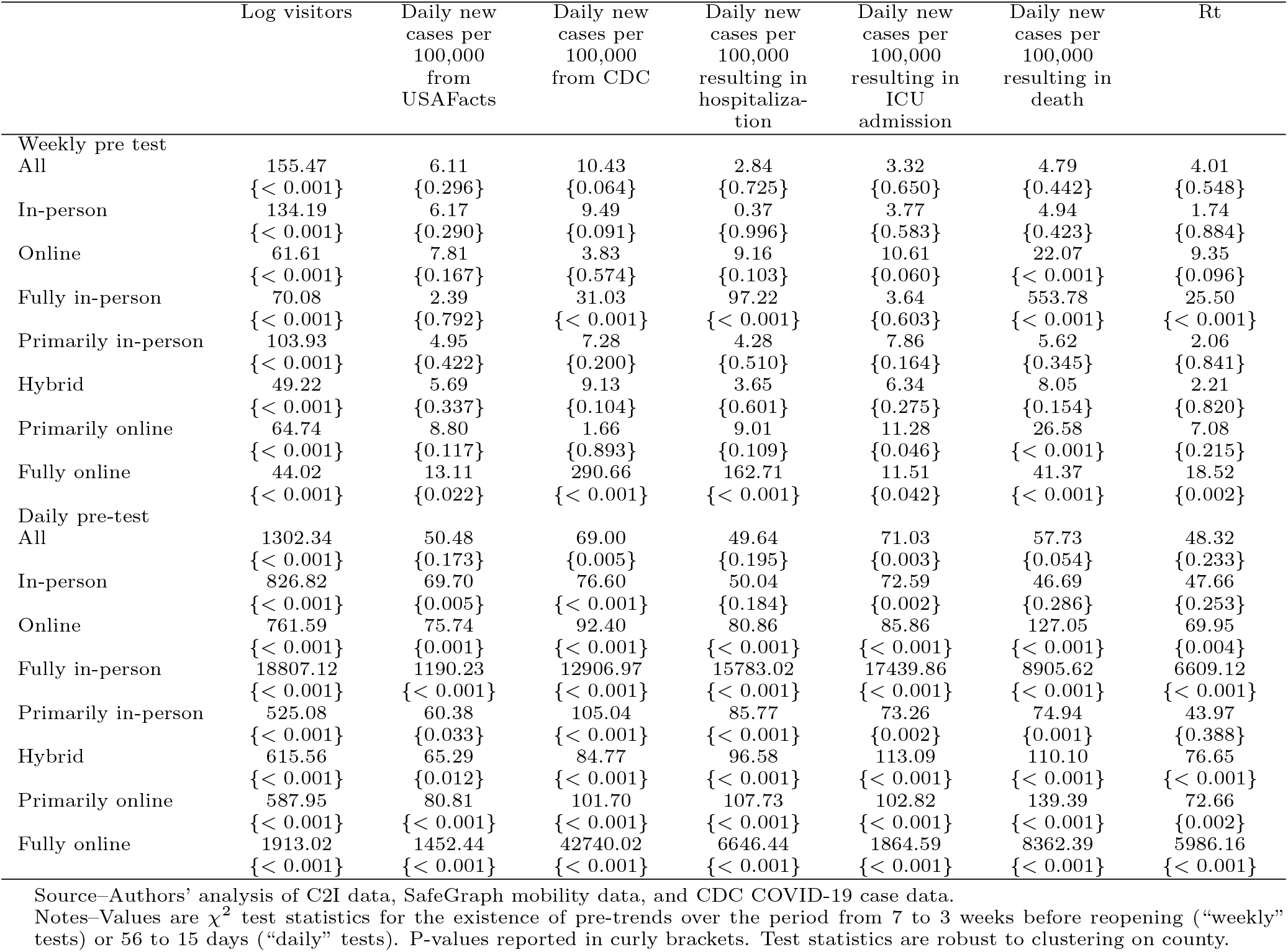
Weekly and daily pre-trend tests

**Table 7:** Teaching modalities across counties

| Teaching modality (# counties) | Coarse modality |  | Fine modality |  |  |  |  |
| --- | --- | --- | --- | --- | --- | --- | --- |
|  | In-person | Online | Fully In Person | Primarily In Person | Hybrid | Primarily Online | Fully Online |
| **Single-Campus counties** |  |  |  |  |  |  |  |
| In-person (393) | 1.00 | 0.00 | 0.08 | 0.58 | 0.35 | 0.00 | 0.00 |
| Online (136) | 0.00 | 1.00 | 0.00 | 0.00 | 0.00 | 0.88 | 0.12 |
| Fully In Person (31) | 1.00 | 0.00 | 1.00 | 0.00 | 0.00 | 0.00 | 0.00 |
| Primarily In Person (226) | 1.00 | 0.00 | 0.00 | 1.00 | 0.00 | 0.00 | 0.00 |
| Hybrid (136) | 1.00 | 0.00 | 0.00 | 0.00 | 1.00 | 0.00 | 0.00 |
| Primarily Online (119) | 0.00 | 1.00 | 0.00 | 0.00 | 0.00 | 1.00 | 0.00 |
| Fully Online (17) | 0.00 | 1.00 | 0.00 | 0.00 | 0.00 | 0.00 | 1.00 |
| **Multi-Campus counties** |  |  |  |  |  |  |  |
| In-person (159) | 0.78 | 0.22 | 0.04 | 0.39 | 0.36 | 0.16 | 0.06 |
| Online (98) | 0.34 | 0.66 | 0.02 | 0.14 | 0.19 | 0.52 | 0.13 |
| Fully In Person (6) | 0.88 | 0.12 | 0.44 | 0.29 | 0.14 | 0.08 | 0.04 |
| Primarily In Person (80) | 0.82 | 0.18 | 0.03 | 0.60 | 0.19 | 0.14 | 0.04 |
| Hybrid (73) | 0.73 | 0.27 | 0.01 | 0.16 | 0.56 | 0.20 | 0.07 |
| Primarily Online (85) | 0.35 | 0.65 | 0.02 | 0.15 | 0.19 | 0.56 | 0.09 |
| Fully Online (13) | 0.28 | 0.72 | 0.01 | 0.08 | 0.19 | 0.30 | 0.42 |

Source—Authors’ analysis of C2I data.
Notes—Values are the row fraction of campuses in each county with the teaching modality indicated by the column header.
Row label indicates the teaching modality assignment of the county.

**Table 8:**
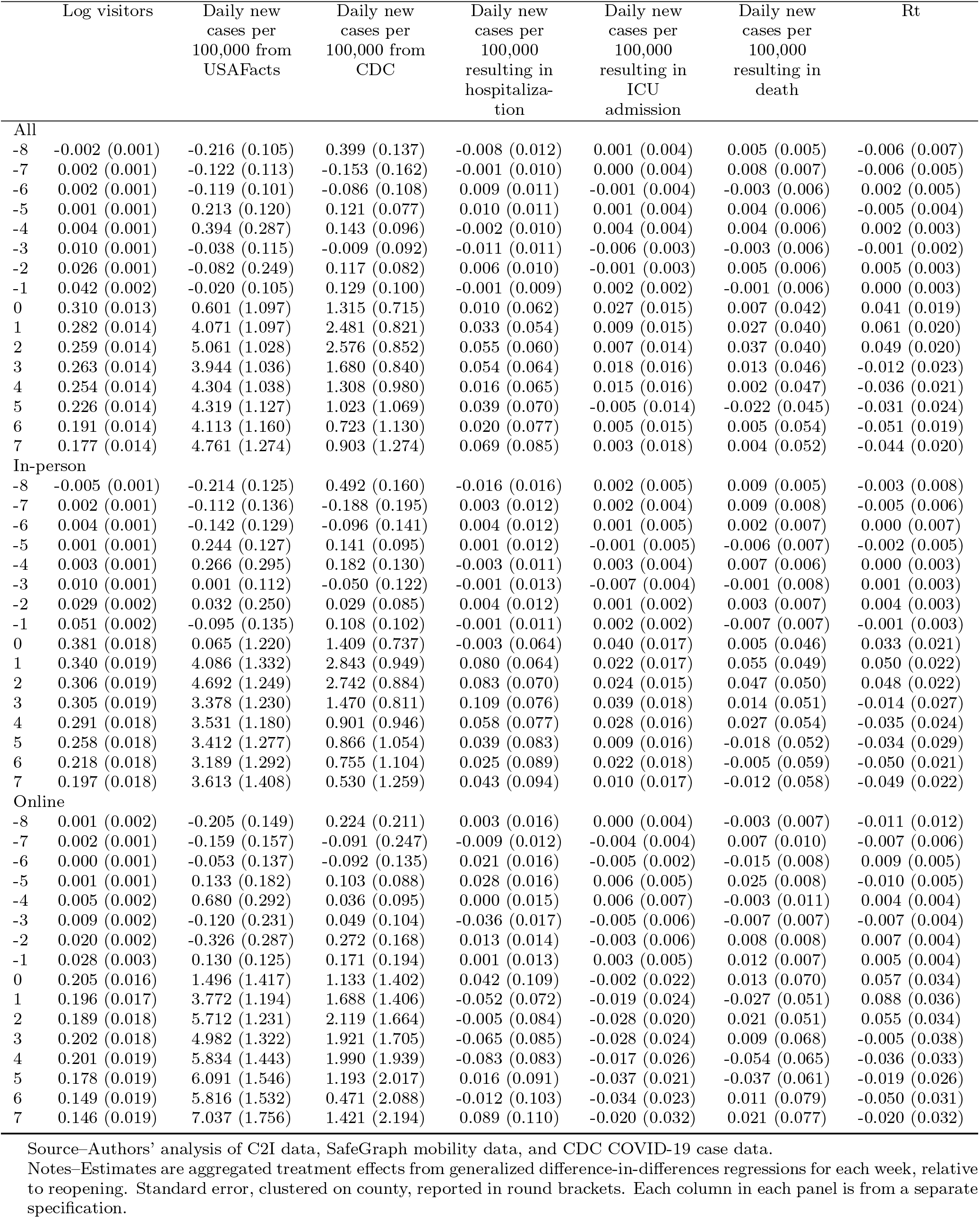
Event study coefficients

## References

1. Chen, M. K., Chevalier, J. A. & Long, E. F. Nursing home staff networks and covid-19. Proc. Natl. Acad. Sci. 118 (2021).

2. Kinner, S. A. et al. Prisons and custodial settings are part of a comprehensive response to covid-19. The Lancet Public Heal. 5, e188–e189 (2020).

3. Richmond, C. S., Sabin, A. P., Jobe, D. A., Lovrich, S. D. & Kenny, P. A. Sars-cov-2 sequencing reveals rapid transmission from college student clusters resulting in morbidity and deaths in vulnerable populations. medRxiv DOI: 10.1101/2020.10.12.20210294 (2020). https://www.medrxiv.org/content/early/2020/10/14/2020.10.12.20210294.full.pdf.

4. Leidner, A. J. Opening of large institutions of higher education and county-level covid-19 incidence—united states, july 6–september 17, 2020. MMWR. Morb. Mortal. Wkly. Rep. 70 (2020).

5. Auger, K. A. et al. Association between statewide school closure and covid-19 incidence and mortality in the us. Jama 324, 859–870 (2020).

6. Laws, R. L. et al. Symptoms and transmission of sars-cov-2 among children—utah and wisconsin, march–may 2020. Pediatrics 147 (2021).

7. Kim, J. et al. Role of children in household transmission of covid-19. Arch. disease childhood (2020).

8. Ludvigsson, J. F. Children are unlikely to be the main drivers of the covid-19 pandemic–a systematic review. Acta Paediatr. 109, 1525–1530 (2020).

9. Mangrum, D. & Niekamp, P. Jue insight: College student travel contributed to local covid-19 spread. J. Urban Econ. 103311, DOI: 10/ghn3cs (2020).

10. Markel, H. et al. Nonpharmaceutical interventions implemented by us cities during the 1918-1919 influenza pandemic. JAMA 298 (2007).

11. Yehya, N., Venkataramani, A. & Harhay, M. O. Statewide Interventions and Covid-19 Mortality in the United States: An Observational Study. Clin. Infect. Dis. DOI: 10.1093/cid/ciaa923 (2020). Ciaa923, https://academic.oup.com/cid/advance-article-pdf/doi/10.1093/cid/ciaa923/33472653/ciaa923.pdf.

12. Marsicano, C. et al. C2i fall 2020 dataset. [data file and code book] (2020).

13. Nierenberg, A. & Pasick, A. Schools briefing: University outbreaks and parental angst. The New York Times (2020).

14. Hubler, S. & Hartocollis, A. How Colleges Became the New Covid Hot Spots. The New York Times (2020).

15. Stubbs, C. W., Springer, M. & Thomas, T. S. The impacts of testing cadence, mode of instruction, and student density on fall 2020 covid-19 rates on campus. medRxiv DOI: 10.1101/2020.12.08.20244574 (2020). https://www.medrxiv.org/content/early/2020/12/09/2020.12.08.20244574.full.pdf.

16. Lu, H. et al. Are college campuses superspreaders? a data-driven modeling study. Comput. Methods Biomech. Biomed. Eng. 0, 1–11, DOI: 10.1080/10255842.2020.1869221 (2021). PMID: 33439055, https://www.tandfonline.com/doi/pdf/10.1080/10255842.2020.1869221.

17. Li, Y. et al. Association of university reopening policies with new confirmed covid-19 cases in the united states. medRxiv DOI: 10.1101/2020.12.11.20247353 (2021). https://www.medrxiv.org/content/early/2021/01/04/2020.12.11.20247353.full.pdf.

18. Badruddoza, S. & Amin, M. D. Causal impacts of teaching modality on us covid-19 spread in fall 2020 semester. medRxiv (2020).

19. Aleta, A., Martín-Corral, D. A., Pastore y Piontti & Moreno, Y. Modelling the impact of testing, contact tracing and household quarantine on second waves of covid-19. Nat. Hum Behav 4, 964–971 (2020).

20. Liu, Q.-H. et al. The covid-19 outbreak in sichuan, china: Epidemiology and impact of interventions. PLOS Comput. Biol. 16, 1–14, DOI: 10.1371/journal.pcbi.1008467 (2021).

21. CDC. Covid-19 vaccine breakthrough infections reported to cdc — united states, january 1–april 30, 2021. mmwr morb mortal wkly rep 2021. MMWR Morb Mortal Wkly Rep 2021 70, 792–793 (2021).

22. Centers for Disease Control and Prevention, COVID-19 Response. Covid-19 case surveillance data access, summary, and limitations (version date: December 31, 2020) (2020).

23. Denny, T. N. et al. Implementation of a pooled surveillance testing program for asymptomatic sars-cov-2 infections on a college campus—duke university, durham, north carolina, august 2–october 11, 2020. Morb. Mortal. Wkly. Rep. 69, 1743 (2020).

24. Paltiel, A. D., Zheng, A. & Walensky, R. P. Assessment of sars-cov-2 screening strategies to permit the safe reopening of college campuses in the united states. JAMA network open 3, e2016818–e2016818 (2020).

25. Goodman-Bacon, A. Difference-in-Differences with Variation in Treatment Timing. Working Paper 25018, National Bureau of Economic Research (2018). DOI: 10.3386/w25018.

26. Wilson, C. The u.s. is entering a new covid-19 vaccination crisis. https://time.com/6046880/covid-19-vaccine-slowdown/ (2021).

27. The New York Times. Coronavirus (covid-19) data in the united states (2021).

28. Moon, S. This california university is telling students to vacate dorms just 1 week after starting classes. https://www.cnn.com/world/live-news/coronavirus-pandemic-08-31-20-intl/index.html (2020).

29. Yan, H., Fox, M. & Gumbrecht, J. CDC official affirms coronavirus deaths really are coronavirus deaths. https://www.cnn.com/2020/09/02/health/us-coronavirus-wednesday/index.html (2020).

30. Diesel, J. Covid-19 vaccination coverage among adults—united states, december 14, 2020–may 22, 2021. MMWR. Morb. Mortal. Wkly. Rep. 70 (2021).

31. Marsicano, C. R., Koo, D. & Rounds, E. G. Covid-19 stats: College and university covid-19 student testing protocols, by mode of instruction (n = 1,849) — united states, spring 2021. CDC Morb. Mortal. Wkly. Rep. 70, 535 (2021).

32. NYT. Coronavirus in the u.s.: Latest map and case count. https://www.nytimes.com/interactive/2021/us/covid-cases.html (2021).

33. Prem, K. et al. Projecting contact matrices in 177 geographical regions: an update and comparison with empirical data for the covid-19 era. medRxiv DOI: 10.1101/2020.07.22.20159772 (2020). https://www.medrxiv.org/content/early/2020/07/28/2020.07.22.20159772.full.pdf.

34. Marsicano, C. C2I: The College Crisis Initiative – Crisis to Innovation. https://collegecrisis.org/ (2020).

35. Department of Homeland Security. Colleges and Universities Campuses (2020).

36. Weill, J. A., Stigler, M., Deschenes, O. & Springborn, M. R. Social distancing responses to COVID-19 emergency declarations strongly differentiated by income. Proc. Natl. Acad. Sci. 117, 19658–19660, DOI: 10/ghcbfn (2020).

37. Holtz, D. et al. Interdependence and the cost of uncoordinated responses to COVID-19. Proc. Natl. Acad. Sci. DOI: 10/gg6rkd (2020).

38. Andersen, M. Early Evidence on Social Distancing in Response to COVID-19 in the United States. SSRN Scholarly Paper ID 3569368, Social Science Research Network, Rochester, NY (2020). DOI: 10.2139/ssrn.3569368.

39. Andersen, M., Maclean, J. C., Pesko, M. F. & Simon, K. I. Effect of a Federal Paid Sick Leave Mandate on Working and Staying at Home: Evidence from Cellular Device Data. Working Paper 27138, National Bureau of Economic Research (2020). DOI: 10.3386/w27138.

40. Gupta, S. et al. Tracking Public and Private Response to the COVID-19 Epidemic: Evidence from State and Local Government Actions. Working Paper 27027, National Bureau of Economic Research (2020). DOI: 10.3386/w27027.

41. Nguyen, T. D. et al. Impacts of State Reopening Policy on Human Mobility. Working Paper 27235, National Bureau of Economic Research (2020). DOI: 10.3386/w27235.

42. Ruggles, S. et al. IPUMS USA: Version 10.0, DOI: 10.18128/D010.V10.0 (2020).

43. Callaway, B. & Sant’Anna, P. H. C. Difference-in-Differences with Multiple Time Periods. SSRN Scholarly Paper ID 3148250, Social Science Research Network, Rochester, NY (2019). DOI: 10.2139/ssrn.3148250.

44. Rambachan, A. & Roth, J. An honest approach to parallel trends. Unpubl. manuscript, Harv. Univ. (2021).

## References

1. Du, Z. et al. Serial interval of covid-19 among publicly reported confirmed cases. Emerg. infectious diseases 26, 1341 (2020).

2. Keeling, M. J. & Rohani, P. Modeling Infectious Diseases in Humans and Animals (Princeton University Press, 2011).

3. Van Kerkhove, M. D., Bento, A. I., Mills, H. L., Ferguson, N. M. & Donnelly, C. A. A review of epidemiological parameters from ebola outbreaks to inform early public health decision-making. Sci. Data 2, 150019, DOI: 10.1038/sdata.2015.19 (2015).

4. Liu, Q.-H. et al. Measurability of the epidemic reproduction number in data-driven contact networks. Proc. Natl. Acad. Sci. 115, 12680–12685, DOI: 10/ggfn24 (2018).

5. Liu, Q.-H. et al. The covid-19 outbreak in sichuan, china: Epidemiology and impact of interventions. PLOS Comput. Biol. 16, 1–14, DOI: 10.1371/journal.pcbi.1008467 (2021).

6. Cori, A., Ferguson, N. M., Fraser, C. & Cauchemez, S. A New Framework and Software to Estimate Time- Varying Reproduction Numbers During Epidemics. Am. J. Epidemiol. 178, 1505–1512, DOI: 10/f5gwtn (2013).

7. Rambachan, A. & Roth, J. An honest approach to parallel trends. Unpubl. manuscript, Harv. Univ. (2021).

